# Purely data-driven exploration of COVID-19 pandemic after three months of the outbreak

**DOI:** 10.1101/2020.04.08.20057638

**Authors:** Shirali Kadyrov, Hayot Berk Saydaliev

## Abstract

It has been three months since the novel coronavirus (COVID-19) pandemic outbreak. Many research studies were carried to understand its epidemiological characteristics in the early phase of the disease outbreak. The current study is yet another contribution to better understand the disease properties by parameter estimation of mathematical SIR epidemic modeling. The authors use Johns Hopkins University’s dataset to estimate the basic reproduction number of COVID-19 for representative countries (Japan, Germany, Italy, France, and Netherlands) selected using cluster analysis. As a by-product, the authors estimate transmission, recovery, and death rates for each selected country and carry statistical tests to see if there are any significant differences.

## Introduction

Officially named as COVID-19, the novel coronavirus pneumonia outbreak was identified first time in Wuhan, China in the late December 2019. It is recognized as a severe respiratory illness similar to MERS-CoV and SARS-CoV. A review article (Fan, Zhao, Shi, & Zhou, 2019) published on March 2, 2019 foresees future SARS- or MERS-like coronavirus disease to originate from bats find it most likely to start in China.

Shortly it was characterized as a pandemic and World Health Organization (WHO) declared it to be “Public Health Emergency of International Concern” on 30 January 2020. As of April 8, 2020, over 1,436,833cases of COVID-19 are reported to WHO, from over 209 countries and territories around the world with more than 82,421 fatalities and about 303,721 recovered.

Obtaining epidemiological characteristics such as basic reproduction number, incubation period, infectious period, and death rate are crucial in better understanding the pandemic outbreak. Shortly after the first statistics made available, various research studies of COVID-19 carried to estimate these parameter values see e.g. (Wu & McGoogan, 2020) (Lai, Shih, Ko, Tang, & Hsueh, 2020). The study (Zhao, et al., 2020) from January 2020 estimates the basic reproduction number *R*_0_ to range from 2.24 (95%CI: 1.96-2.55) to 5.71 (95%CI: 4.24-7.54) using *R*_0_ *= 1/M*(−*γ*) formula where *M* is the moment generating function for the serial interval of the COVID-19 and *γ* is the intrinsic growth rate. Another early work (Read, Bridgen, Cummings, Ho, & Jewell, 2020) estimates R_0_ to be 3.11 (95%CI, 2.39-4.13) using deterministic SEIR metapopulation model. The same work estimates the transmission rate *β*, to be 1.94 (95%CI, 1.25–6.71) and infectious period to be 1.61 days (95%CI, 0.35–3.23) within Wuhan, China. The review of twelve recent works shows that R_0_ ranges from 1.4 to 6.49, with a mean of 3.28, a median of 2.79 and interquartile range (IQR) of 1.16 (Liu, Gayle, Wilder-Smith, & Rocklöv, 2020). While early reports of Chinese Center for Disease Control and Prevention (CDC) suggest the infectious period to be 9 days, another recent work (You, et al., 2020) reports the mean infectious period to be 10.91 days (SD=3.95).

For various mathematical models formulated to forecasting the development of the disease and estimating the parameters we refer to (Rabajante, 2020) and references therein.

Our goal in this article is to estimate death rates for COVID-19 for selected representative countries and use these parameters to estimate recovery rates and basic reproduction numbers from a deterministic mathematical model. We then compare these results to see if there are any significant differences among countries.

This rest of the paper is organized as follows, we introduce the methodology including data, the mathematical model, and parameter estimation technique. Then comes the results section where we report our findings. Finally, we end with discussion section where we interpret the findings and provide recommendations for future research.

## Method

### Data Set

This study uses Johns Hopkins University’s COVID-19 data made available at GitHub repository (JHU, 2020). The dataset includes confirmed, recovered, and death cases for almost all countries of the world over the period of January 22, 2019 to March 23, 2020 on a daily basis. The epidemiological characteristics of COVID-19 in the USA, South Korea, Italy, France, China, and Iran are studied and this list of selected countries is based on cluster analysis.

### Theoretical Model

Let *N* be the total population of a country. The following diagram summarizes the disease transmission process.

**Figure 1a.**
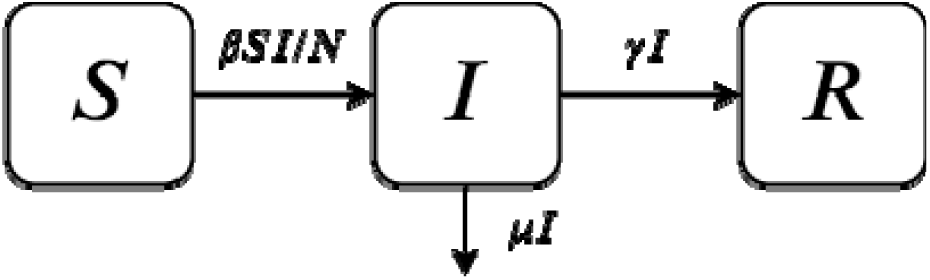
Model diagram

The corresponding classical deterministic *SIR* epidemic model is given by

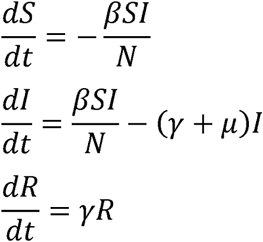

where, *S, I, R* are Susceptible, Infectious, and Recovered classes and parameters *β, γ, µ* are transmission, recovery, and death rates, respectively. Since we have a dataset given in a daily basis, we fix our time units in terms of days. For this model, the basic reproduction number is given by the formula

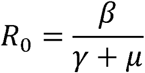

We note that this is a system of autonomous nonlinear ordinary differential equations. The nonlinearity term *SI* is due to law of mass-action, which is the reason for the absence of non-trivial closed form. However, computer assisted numerical approximations are available. There are variations of this model where one may include new classes such as exposed and quarantine and may also consider demographic characteristics such as birth and natural death rates etc. As our parameter estimations well fit to the current model, we decided to keep it as is.

### Empirical Model Specification

#### Clustering

Before applying the statistics model (Least square) and SIR methods, this study verifies the cluster analysis. As reported above, there are over 184 countries where the COVID-19 outbreak occurred so far. Analyzing each country separately and reporting results in a research article is not very convenient. Therefore, we decided to run cluster analysis to first group the countries according to similarity of confirmed cases and then select one representative country from each cluster. Here, cluster analysis carried using country recovery rates (***γ***) and the country doubling periods of the confirmed cases. The last equation ***SIR*** model reads 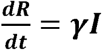, which can be rewritten in the discretized version as

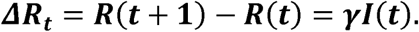

Both ***ΔR*** and ***I***(***t***) can be computed from the dataset and ***γ*** can be obtained as the slope of the equation. Before the confirmed case gets very large, one can safely assume that ***S ≈ N*** which together with the second equation of ***SIR*** gives 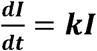 with the growth rates is ***k*** *=* ***β*** − ***µ*** − ***γ***. This has a solution ***I***(***t***) *=* ***I***(***0***) ***e***^***kt***^ from which one can obtain

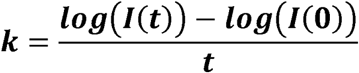

where we take time ***t*** to be the last day available in the dataset and ***t*** *=* **0** is taken so that ***I***(**0**) is nonzero. We warn here that ***I***(***t***)in the model is the total number of infectious at time ***t*** which is different from total confirmed cases at time ***t*.** However, if ***I*** has exponential growth then so does the total confirmed cases. Once the growth rate is estimated for each country, the doubling period is computed to be (***ln* 2)***/****k***.

We conduct ***k***-means cluster analysis which is sensitive to outliers. Hence, we first cleared outliers from our data using box plot analysis, then used the elbow method to determine the number of centroids. Finally, the cluster analysis is carried for the normalized variables, z-scores, and one representative is selected from each cluster. Moreover, we did not completely avoid the outliers, instead selected two representative countries one with too high recovery rate and one with too high doubling period. Once the list of representative countries are selected, we turn to parameter estimation and prediction.

#### Parameter estimation

To estimate the parameters *β, γ, µ* for the *SIR* system one usually considers nonlinear least square analysis. However, models are simplified versions of real life systems and are not always behave well with parameter estimations. What we will do is to estimate *γ, µ* parameters using simple linear regression. Then, use these point estimates in the SIR model to estimate *β.* More specifically, we use ‘scipy.integrate.ode’ function of python programming language to simulate S(t), I(t), and R(t) where we let initial values as S(0)=country population, I(0)= the first observed number of cases in the country, and R(0)=0. Then, we call ‘scipy.optimize.curve_fit’ function for least-square fitting of the theoretical model solution to the observed daily number of confirmed cases and recovered individuals.

#### Analysis and results

The countries with fewer data resulted in unrealistic doubling periods and/or almost zero recovering rates, these countries were eliminated. Eliminating countries with doubling period more than 10 resulted in 152 countries left. On the other hand, if we eliminate those countries with very small (<0.0001) recovery rate we are left with only 67 countries. The eliminated countries are those with few data reports or with missing data for recovered individuals.

Further remaining outliers were cleared using box plot analysis which resulted in 54 countries left. The final box plot results for the 54 countries are shown in Figure 1.

**Figure 1.**
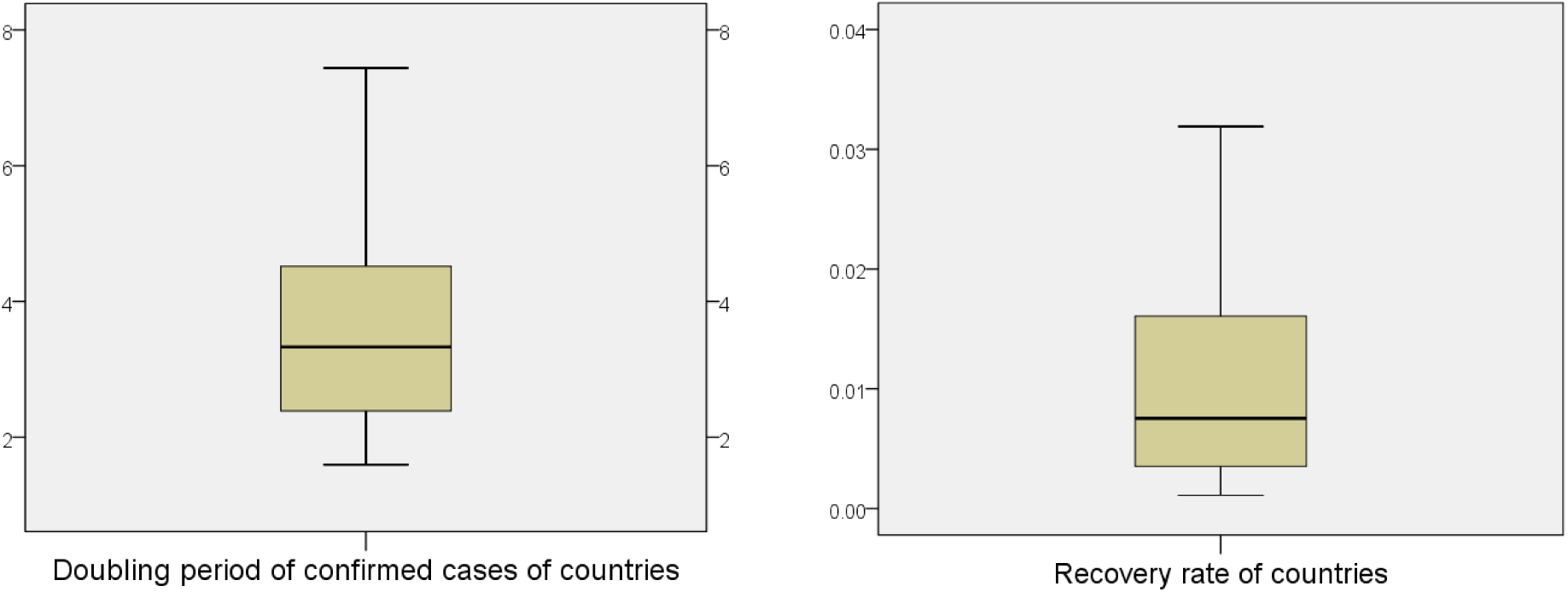
Box plots of recovery rates and case doubling periods

In Figure 2, we see the distributions of both doubling periods and recovery rates. In particular, we see that the mean doubling period for 54 countries is 3.60 (95%CI: 3.22-3.99).

**Figure 2.**
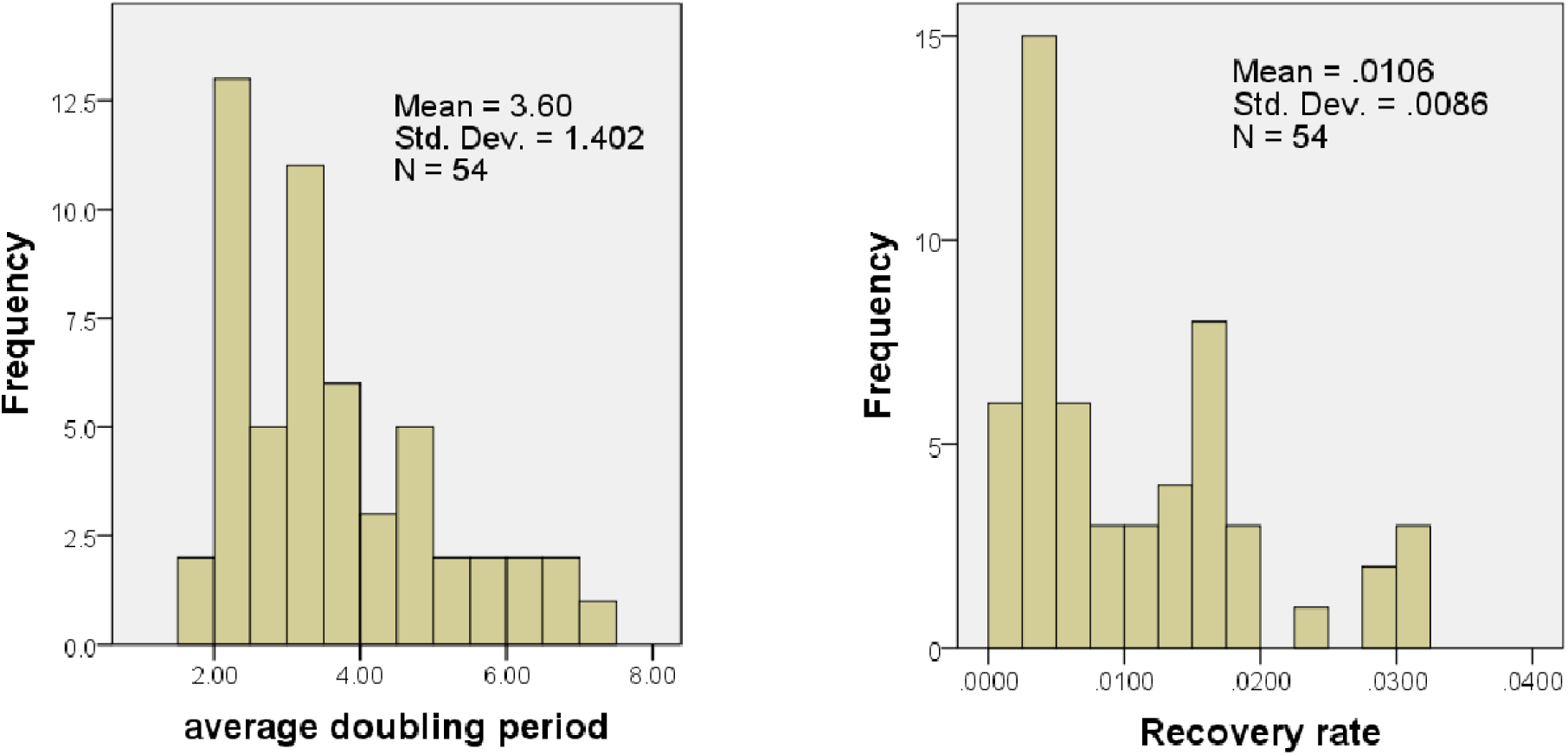
Distribution of doubling period for the confirmed cases and recovery rates

Elbow method summary is shown in Figure 3. Here we see that there are five points with sum of squared distances more than 1. However, since the total remaining countries are only 54 we decided to fix the number of clusters to 4.

**Figure 3.**
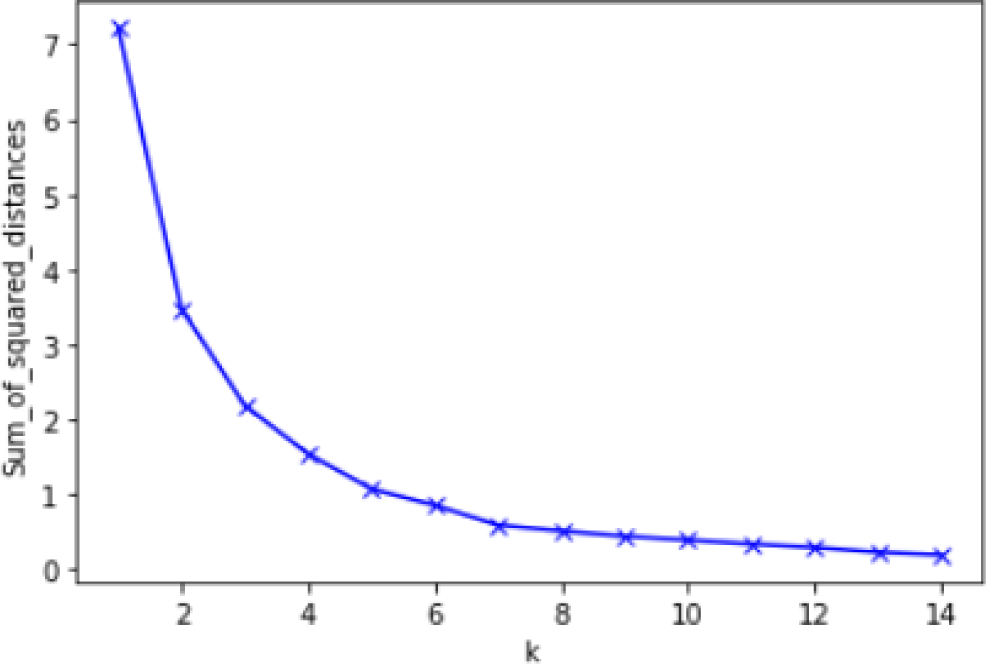
Elbow method For Optimal k

The bar charts of the z-scores of the variables for each cluster is shown in Figure 4 and the list of countries in each cluster is given in Table 1. Both recovery rate and doubling period are found to be significant predictors with *F*(3, 50) = 76.19, *p* <0.001 and *F*(3, 50) = 45.17, *p* <0.001 respectively.

**Table 1.** Countries in each Cluster.

|  |  |  |  |
| --- | --- | --- | --- |
| Cluster | 1 | 14 | Afghanistan, Australia, Cruise Ship, Egypt, Georgia, India, Japan, South Korea, Malaysia, Monaco, Nigeria, Russia, Singapore, United Arab Emirates |
|  | 2 | 25 | Albania, Armenia, Bangladesh, Brunei, Bulgaria, Chile, Colombia, Costa Rica, Côte d'Ivoire, Croatia, Cyprus, Germany, Greece, Israel, Lebanon, Luxembourg, Morocco, Poland, San Marino, Saudi Arabia, Slovakia, Switzerland, Trinidad and Tobago, Ukraine, US |
|  | 3 | 10 | Azerbaijan, Belgium, Burkina Faso, Iceland, Indonesia, Italy, Jamaica, Kuwait, Senegal, Spain |
|  | 4 | 5 | Belarus, France, Hungary, Iraq, Pakistan |

**Figure 4.**
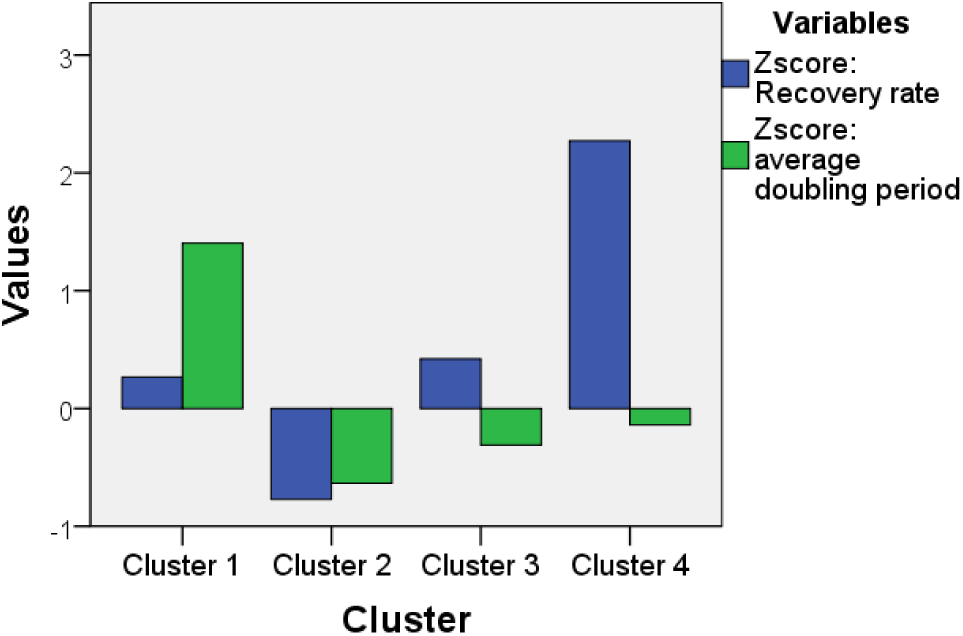
Final Cluster Centers

We see that the first cluster contains 14 countries with high doubling period, the second cluster contains 25 countries with low recovery rate and low doubling period, the third includes 10 countries with somewhat average recovery rate and average doubling period, and the last cluster has 5 countries with high recovery rate.

From each cluster we select one representative country, they are Japan, Germany, Italy, and France. We select one more country as a representative of outliers, namely Netherland with a low case doubling period (2.08 days) and too low recovery period (<0.001).

For the selected countries, least square parameter estimation using theoretical SIR model is carried and results are summarized in Table 2.

**Table 2.** Summary of parameter estimations with standard deviations in brackets

|  | Japan | Germany | Italy | France | Netherlands |
| --- | --- | --- | --- | --- | --- |
| Transmission | 0.1898 | 0.1895 | 0.3481 | 0.2022 | 0.4017 |
| rate ($\beta$) | (1.65e-05) | (3.93e-06) | (1.64e-05) | (4.34e-06) | (0.0003) |
| Recovery rate | 0.0284 | 0.0018 | 0.027 | 0.0061 | 0.0002 |
| ( ) | (1.72e-05) | (3.89e-06) | (1.66e-05) | (4.32e-06) | (0.00029) |
| Death rate ( ) | 0.0426 | 0.0035 | 0.1202 | 0.042 | 0.0472 |
| R0 | 2.67 | 35.74 | 2.36 | 4.20 | 8.47 |
| Case doubling period | 6.71 | 3.84 | 3.28 | 4.22 | 2.08 |
| Mean error | 7 | 45 | 190 | 36 | 45 |

As mentioned in the methodology section, the death rates were estimated using linear regression which is then substituted into model to estimate both transmission rates and recovery rates. The reported mean error is computed as the L2-norm of residuals divided by the total data points used in the estimation. Figure 5 shows the plot of data points vs. the simulation with estimated parameter values for each representative country. The three hundred day simulations of *SIR* model after the start of the outbreak are provided in the appendix.

**Figure 5.**
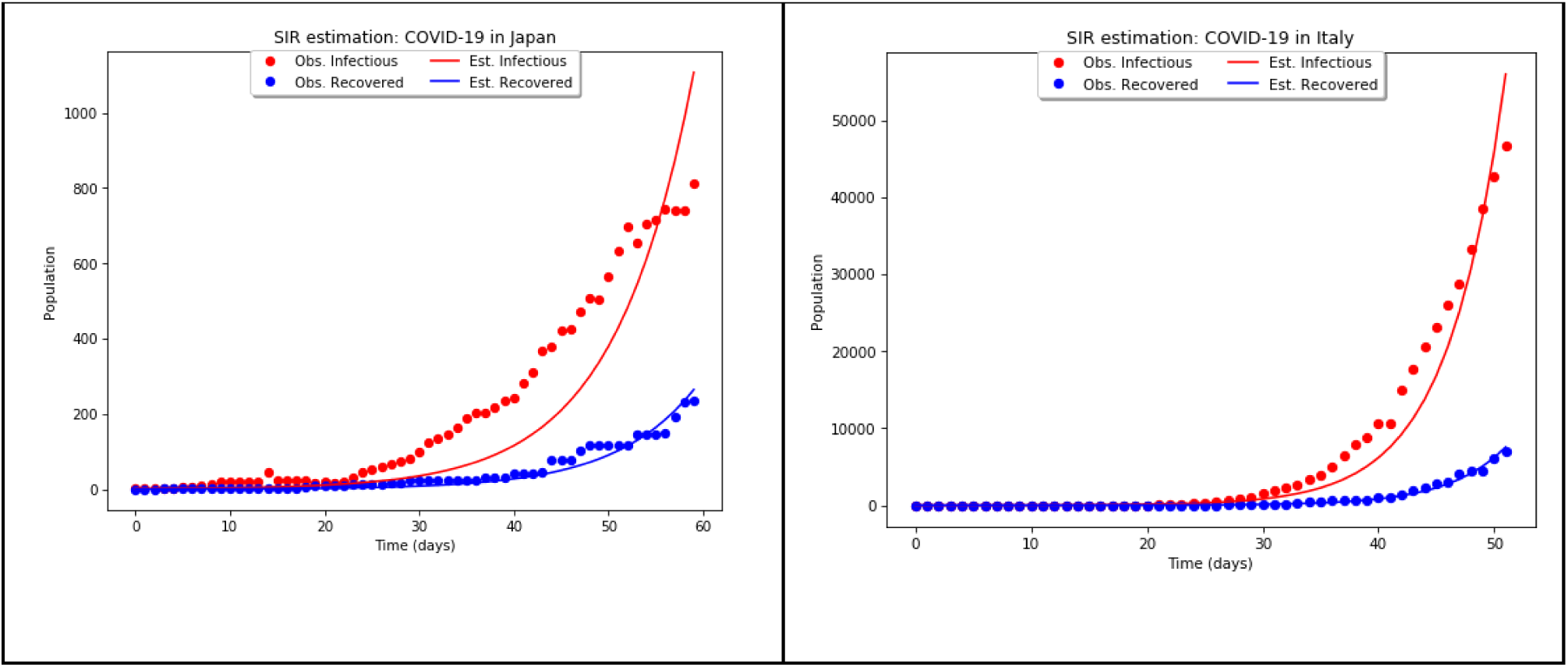
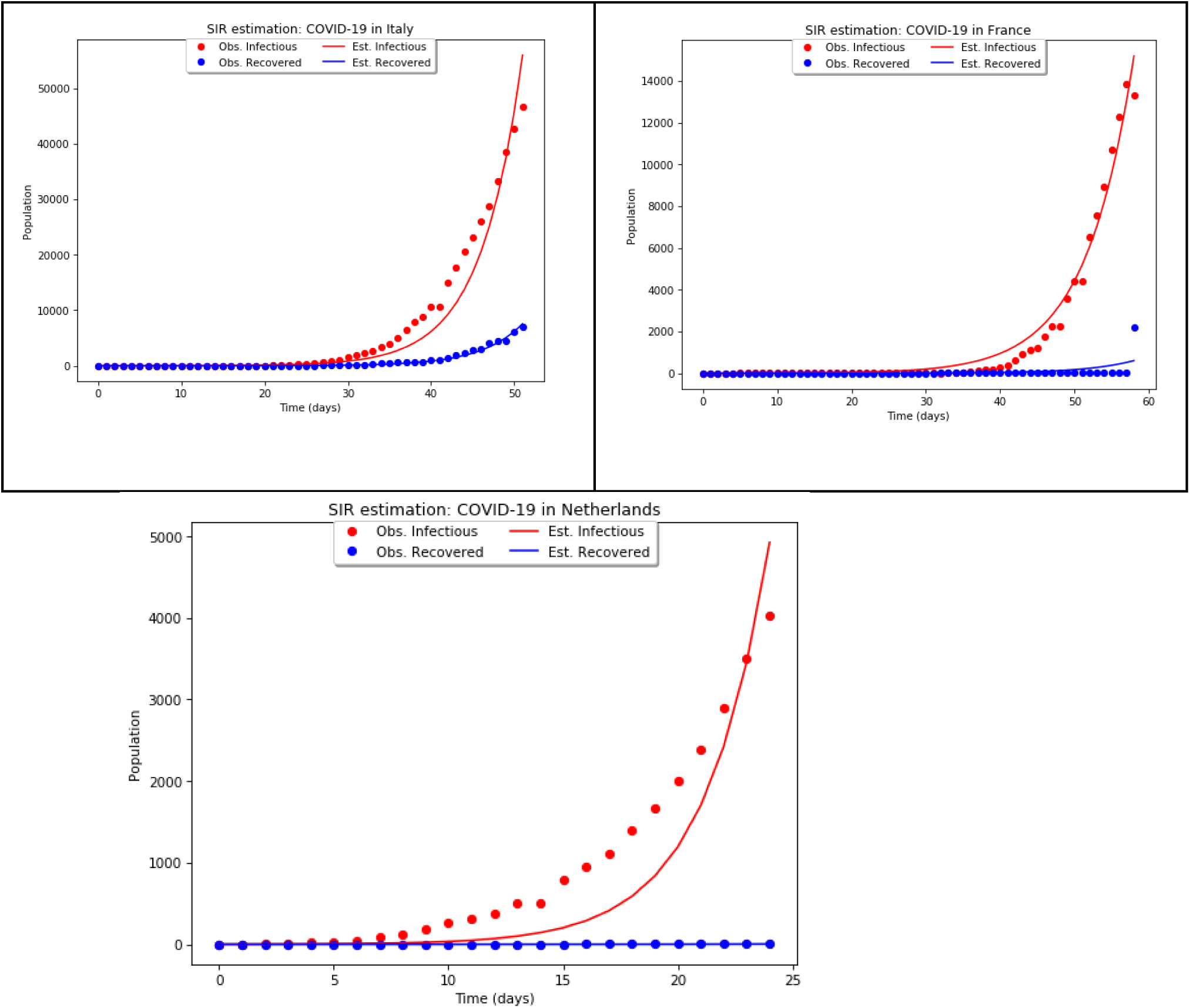
Plot of the data (dotted curve) vs. simulation (solid curve). The red and blue colors correspond to infectious and recovered cases respectively.

## Discussion and Conclusion

In this article we considered The Kermack and McKendrick SIR model to estimate COVID-19 epidemiological characteristics using Johns Hopkins University dataset over the period from January 22 to March 22. Our approach is purely data-driven without relying any parameters reported before. 183 countries were divided into 5 clusters classifies according to recovery rate and case doubling periods and one country was selected from each cluster as a representative. The summary of parameter estimation results are given in Table 2. Here we see that the death rate varies from 0.0035 to 0.1202 with least being Germany and the highest being Italy. One may expect the similar estimates in within the respective clusters. The reason for such a significant difference in death rates requires further investigation.

When it comes to parameter estimation using nonlinear epidemic models the recovery rate *γ* is usually taken to be the inverse of infectious period. However, estimation results reveal, see Table 2, that this rate is varies from 0.0018 to 0.0284 for the representative countries of four clusters. Taking the inverses yield the range from 35 days to 555 days which is much higher than the infectious period reported before, see.e.g. (CDC), (You, et al., 2020). This indicates that recovery rate should not be taken as the inverse of infectious period provided the dataset (JHU, 2020) is accurate.

As for the basic reproduction number *R*_0_, we have 2.36, 2.67, 4.20, 8.47, and 35.74 for the countries Italy, Japan, France, Netherlands, and Germany respectively. While the first four estimates are in line with earlier reports of *R_*0 for COVID-19, we find *R_*0 estimate of 35.74 for Germany very high. This is consistent with the Germany low death rate of 0.0035 found as death rate appears in the reciprocal of basic reproduction formula.

One of the limitations of the study is the model, SIR, being theoretical. As a theoretical model, the forecasting of the pandemic progression from SIR might be misleading. For our five representative countries we provided the three hundred day simulation in the appendix. What we see from the simulations that the COVID-19 pandemic is more likely to start slowing down within 100-150 days starting from January 22. Looking at the times series of Susceptible individuals we see that over 80% of countries will be affected by the pandemic. However, we note that our model ignores any kinds of preventative situations and as such it is very likely that the total affected case will be much lower. It is possible to consider various improvements of the model where one can include other compartments such as Exposed or Quarantined individuals and consider non-autonomous system where the transmission rate is controlled according to the public awareness such as social distancing, self-quarantines, wearing masks etc.

## Data Availability

Johns Hopkins University dataset is used

## Appendix

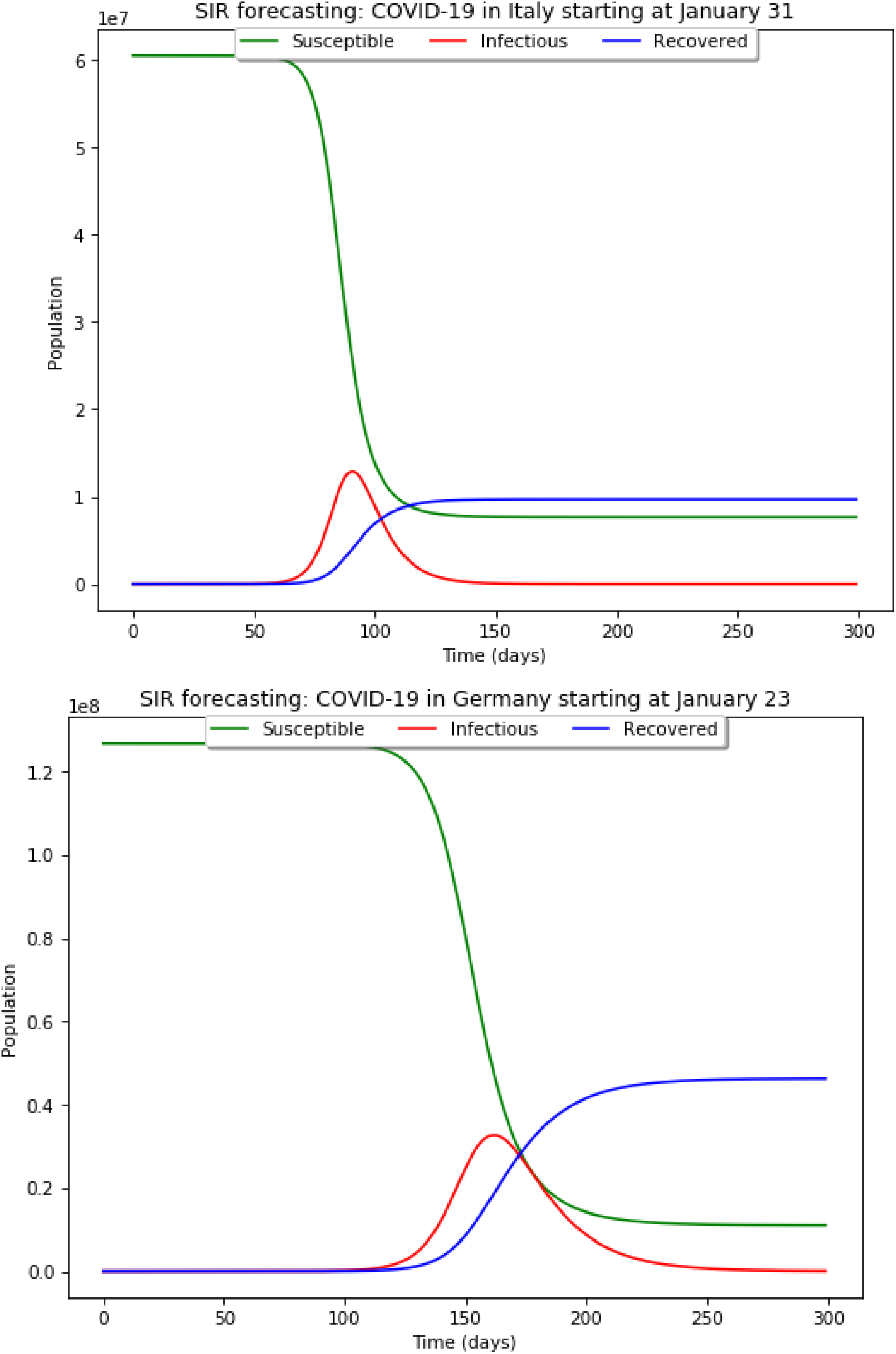

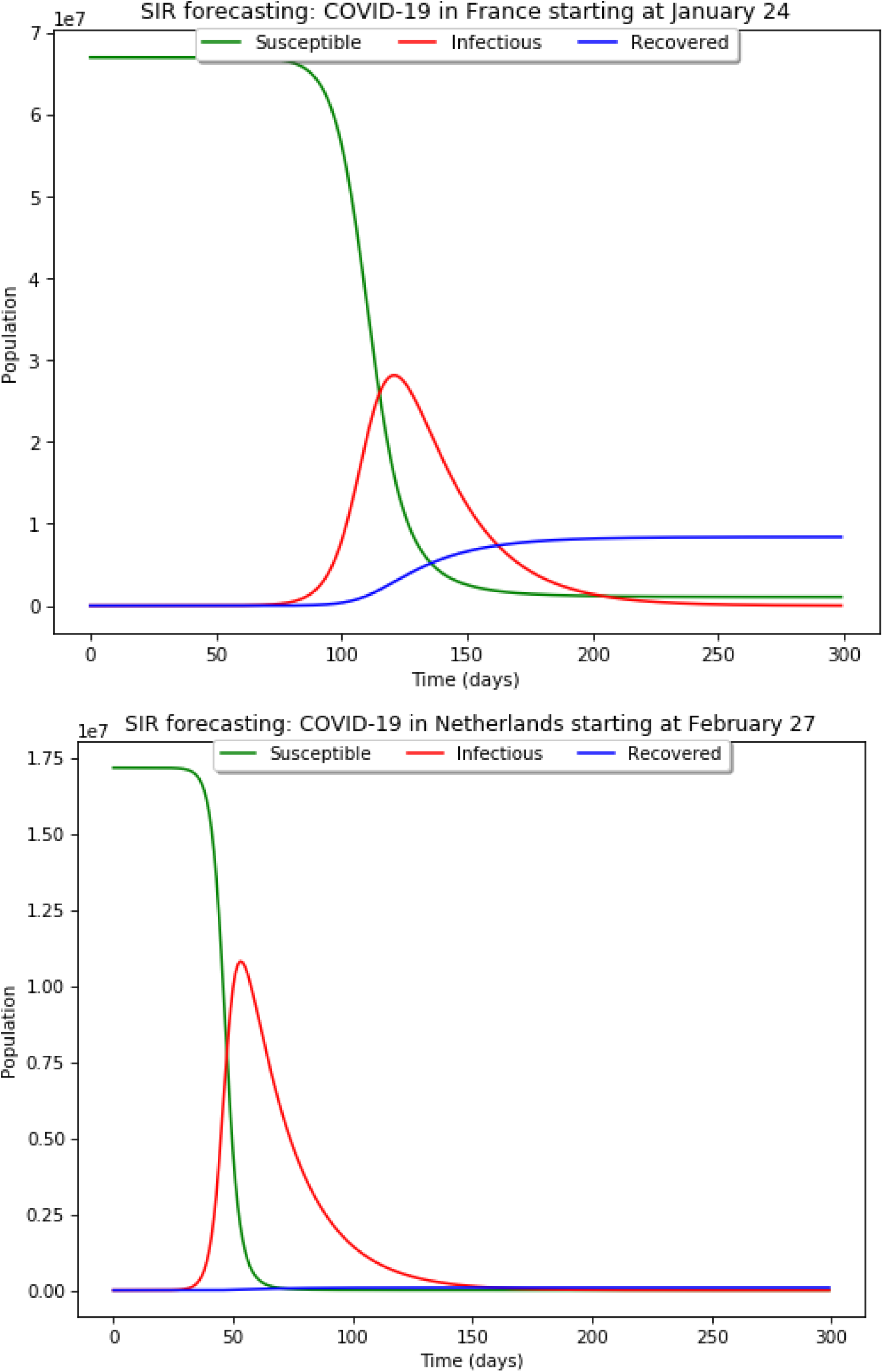

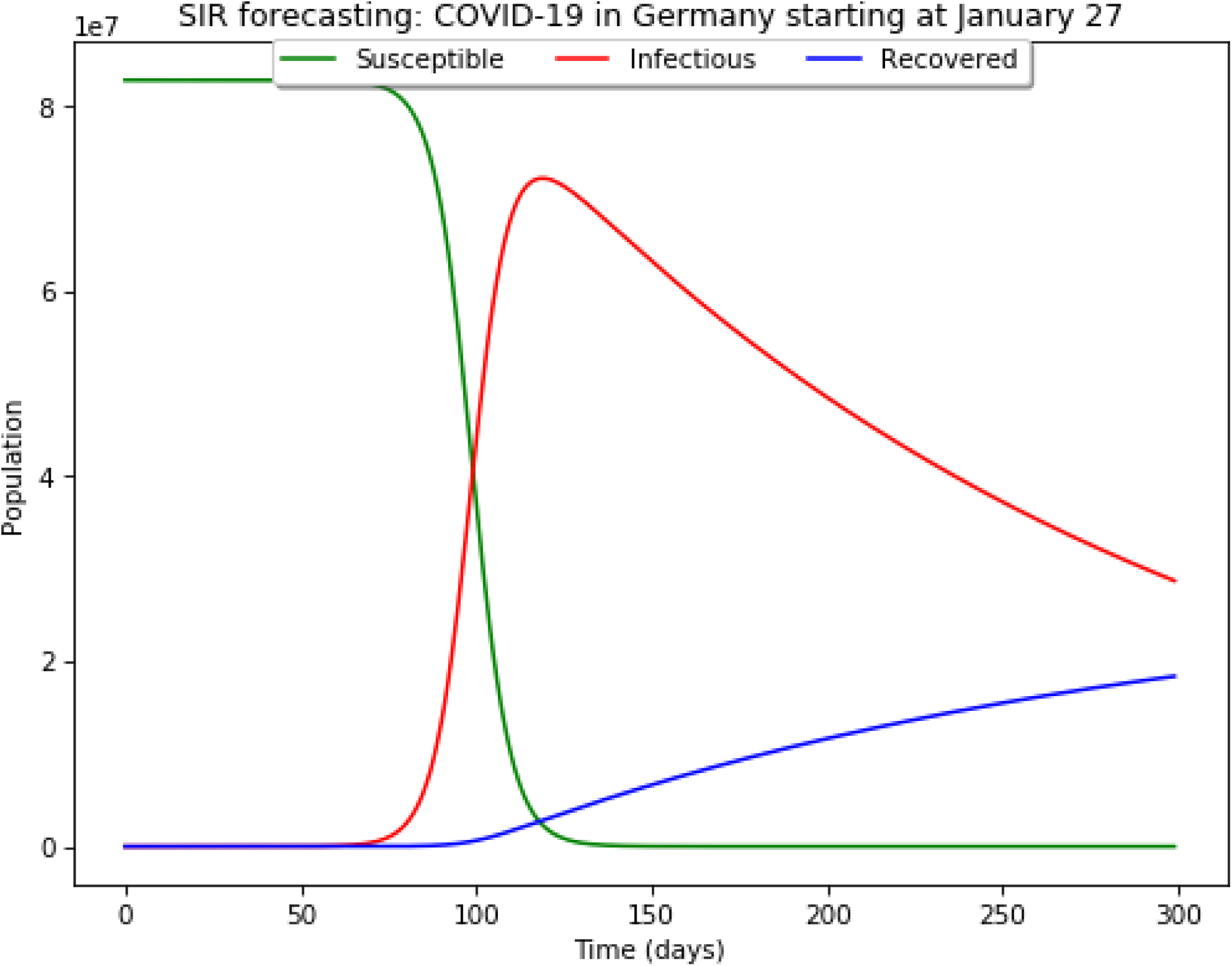

## Bibliography

CDC. (n.d.). Retrieved January 26, 2020, from Chinese Center for Disease Control and Prevention. The 2019 novel coronavirus.: http://www.chinacdc.cn/jkzt/crb/zl/szkb_11803/

Fan, Y., Zhao, K., Shi, Z. L., & Zhou, P. (2019, March 2). Bat Coronaviruses in China. Viruses, 11(3), 210.

JHU. (2020). Novel Coronavirus (COVID-19) Cases, provided by JHU CSSE. Retrieved 2020, from https://github.com/CSSEGISandData/COVID-19: https://github.com/CSSEGISandData/COVID-19

Lai, C.-C., Shih, T.-P., Ko, W.-C., Tang, H.-J., & Hsueh, P.-R. (2020). Severe acute respiratory syndrome coronavirus 2 (SARS-CoV-2) and corona virus disease-2019 (COVID-19): the epidemic and the challenges. 105924.

Liu, Y., Gayle, A., Wilder-Smith, A., & Rocklöv, J. (2020). The reproductive number of COVID-19 is higher compared to SARS coronavirus.

Rabajante, J. (2020). Insights from early mathematical models of 2019-nCoV acute respiratory disease (COVID-19) dynamics. Insights from early mathematical models of 2019-nCoV acute respiratory disease (COVID-19) dynamics.

Read, J. M., Bridgen, J. R., Cummings, D. A., Ho, A., & Jewell, C. P. (2020). Novel coronavirus 2019-nCoV: early estimation of epidemiological parameters and epidemic predictions. medRxiv.

Wu, Z., & McGoogan, J. (2020). Characteristics of and important lessons from the coronavirus disease 2019 (COVID-19) outbreak in China: summary of a report of 72 314 cases from the Chinese Center for Disease Control and Prevention.

You, C., Deng, Y., Hu, W., Sun, J., Lin, Q., Zhou, F., & Zhou, X. H. (2020). Estimation of the Time-Varying Reproduction Number of COVID-19 Outbreak in China. Available at SSRN 3539694.

Zhao, S., Lin, Q., Ran, J., Musa, S. S., Yang, G., Wang, W., & Wang, M. H. (2020). Preliminary estimation of the basic reproduction number of novel coronavirus (2019-nCoV) in China, from 2019 to 2020: A data-driven analysis in the early phase of the outbreak. International Journal of Infectious Diseases, 92, 214–217.

